# A Bayesian Framework for Estimating the Risk Ratio of Hospitalization for People with Comorbidity Infected by the SARS-CoV-2 Virus

**DOI:** 10.1101/2020.07.25.20162131

**Authors:** Xiang Gao, Qunfeng Dong

## Abstract

Estimating the hospitalization risk for people with certain comorbidities infected by the SARS-CoV-2 virus is important for developing public health policies and guidance based on risk stratification. Traditional biostatistical methods require knowing both the number of infected people who were hospitalized and the number of infected people who were not hospitalized. However, the latter may be undercounted, as it is limited to only those who were tested for viral infection. In addition, comorbidity information for people not hospitalized may not always be readily available for traditional biostatistical analyses. To overcome these limitations, we developed a Bayesian approach that only requires the observed frequency of comorbidities in COVID-19 patients in hospitals and the prevalence of comorbidities in the general population. By applying our approach to two different large-scale datasets in the U.S., our results consistently indicated that cardiovascular diseases carried the highest hospitalization risk for COVID-19 patients, followed by diabetes, chronic respiratory disease, hypertension, and obesity, respectively.

**Significance Statement:** We developed a novel Bayesian approach to estimate the hospitalization risk for people with comorbidities infected with the SARS-CoV-2 virus. Our results indicated that cardiovascular diseases carried the highest hospitalization risk for COVID-19 patients, followed by diabetes, chronic respiratory disease, hypertension, and obesity, respectively.

## Introduction

Recently published data from China^1^, Italy^2^, and the U.S.^3^ showed that a large portion of hospitalized COVID-19 patients had at least one preexisting comorbidity or medical condition. Estimating the hospitalization risk for people with a certain comorbidity condition infected by the SARS-CoV-2 virus would be important for developing public health policies and guidance (e.g., planning medical resources) based on risk stratification.

To provide such an estimation, traditional biostatistical methods (e.g., logistic regression) require knowing both the number of infected people who were hospitalized and the number of infected people who were not hospitalized. Although the number of infected people who were hospitalized might be tallied from hospital records, the number of infected people who were not hospitalized may be difficult to estimate. The reason is that the official count of infected people who were not hospitalized is limited to those tested for the virus. Since not everyone infected by the virus is tested, the number of infected people not hospitalized may be undercounted in the traditional analysis. In addition, comorbidity information for people not hospitalized may not always be readily available for traditional biostatistical analyses. For example, Petrilli et al^4^ recently applied multiple logistic regression to identify factors associated with hospital admission using a prospective cohort, but they had stated two main limitations in their study regarding the patients who were not hospitalized: (i) many confirmed patients might not have had their detailed medical history recorded for the study, and (ii) many potential patients might have been omitted from the study since they were not tested for the virus.

To overcome these limitations, we developed a Bayesian approach to estimate the risk ratio of hospitalization for COVID-19 patients with comorbidities. The risk ratio is defined as a ratio of the probability of hospitalization for the infected people with a particular comorbidity (e.g., diabetes) versus the probability of hospitalization for the infected people without any comorbidities. In other words, people without any comorbidities are used as reference for the risk ratio estimation. Our Bayesian approach only needs (i) the number of hospitalized COVID-19 patients and their comorbidity information, which can be reliably obtained using hospital records, and (ii) the prevalence of the comorbidity of interest in the general population, which is regularly documented by public health agencies for common medical conditions (e.g., hypertension, obesity, and diabetes). By applying our approach to two different large-scale datasets in the U.S., we obtained consistent results showing that cardiovascular disease had the highest elevated risk of hospitalization, followed by diabetes, chronic respiratory diseases, hypertension, and obesity, respectively.

## Results

### Estimation with the COVID-NET data

We applied our Bayesian approach to a dataset^5^ of hospitalized COVID-19 patients from the COVID-NET (n=2491). Table 1 provides the summary statistics (median, central 95% Bayesian credible interval) for the estimated posterior distributions of the risk ratio of hospitalization for COVID-19 patients with cardiovascular disease (6.9, 5.1-9.3), diabetes (3.6, 2.9-4.4), COPD (2.6, 2.1-3.2), asthma (2.3, 1.9-2.9), hypertension (1.7, 1.4-2.0), and obesity (1.6, 1.3-1.9). Figure 1A depicts the posterior distributions for these estimated risk ratios.

**Table 1.**
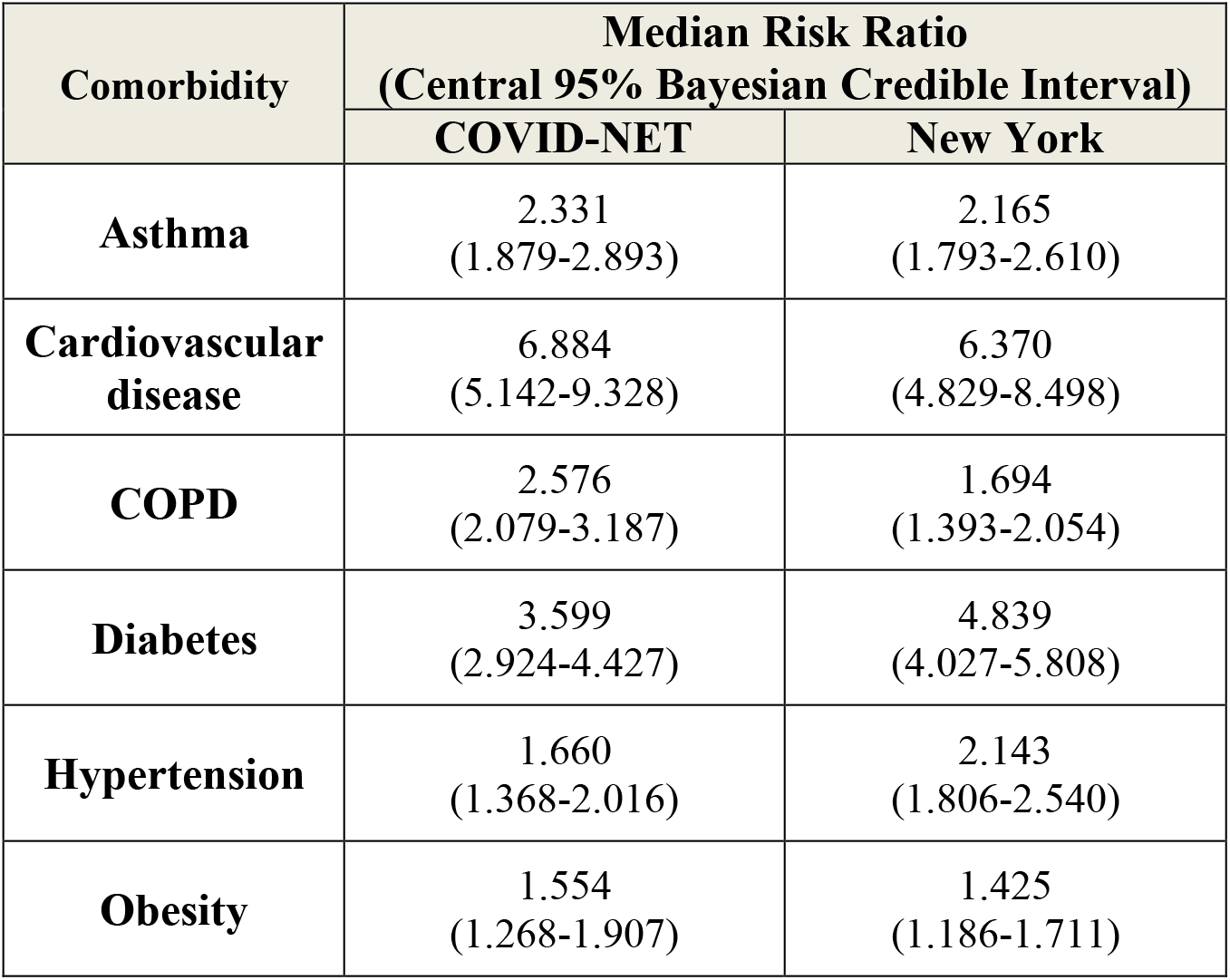
Summary statistics of the posterior distributions of the hospitalization risk for COVID-19 patients with comorbidities

**Figure 1.**
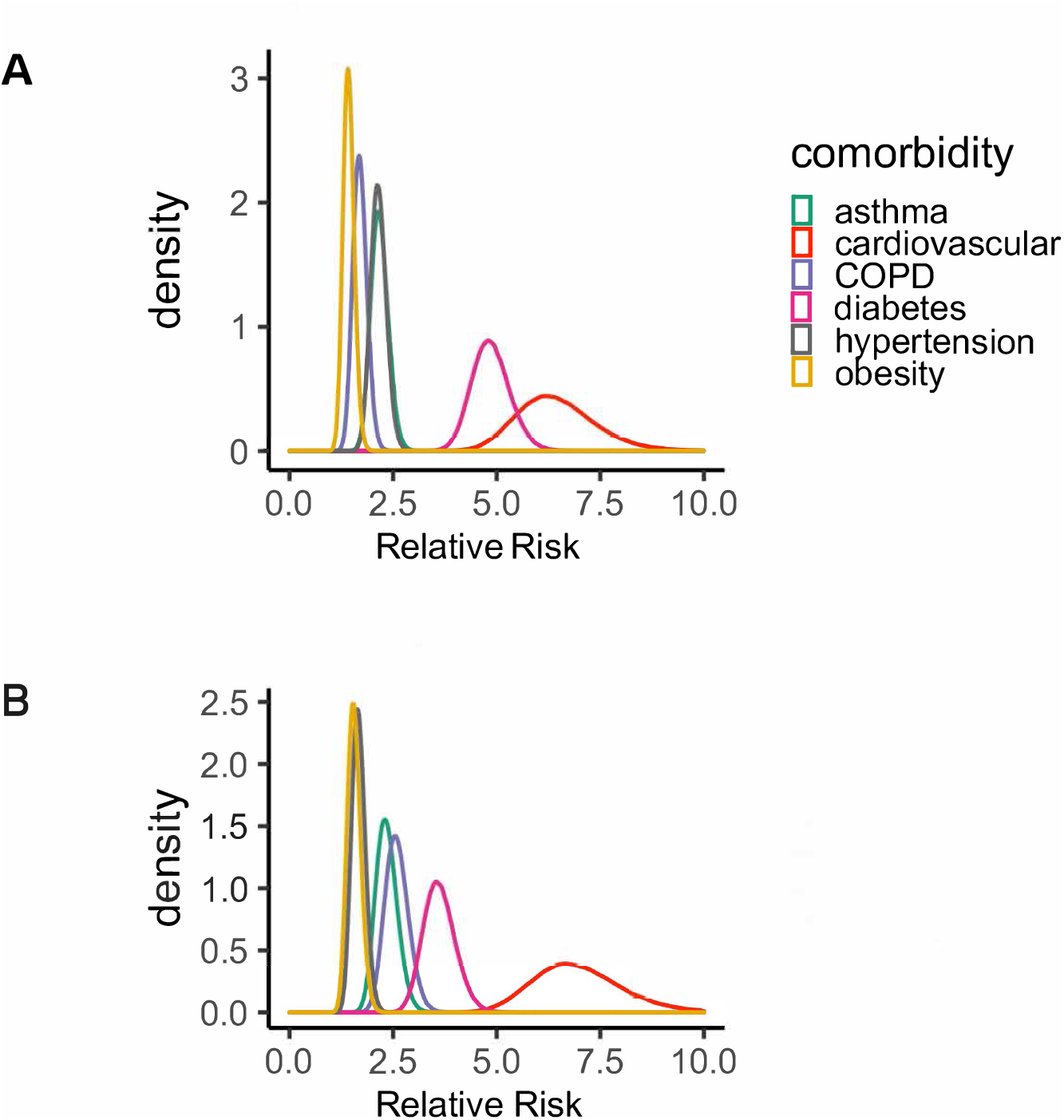
The posterior probability density of the risk ratio of hospitalization for each comorbidity estimated from the datasets of (**A**) COVID-NET and (**B**) New York.

### Estimation with the New York data

A different dataset^3^ from the state of New York (n=5700) was used as a comparison. The results from the New York dataset were similar to the ones from the COVID-NET dataset, showing that cardiovascular diseases (6.4, 4.9-8.5) and diabetes (4.8, 4.0-5.8) significantly increased the risk of hospitalization for COVID-19 patients, followed by asthma (2.2, 1.8-2.6), hypertension (2.1, 1.8-2.5), COPD (1.7, 1.4-2.1), and obesity (1.4, 1.2-1.7) (Table 1, fig. 1B). The only major difference in the ranking of the risk from the COVID-NET and the New York datasets was for COPD, with a lower risk estimated from the New York dataset compared to the COVID-NET dataset.

## Discussion

Using two different large-scale datasets from COVID-NET and New York, our approach obtained similar results, which strongly indicated that COVID-19 patients with cardiovascular disease or diabetes had the highest elevated risk of hospitalization. For example, the risk ratio of hospitalization for COVID-19 patients with cardiovascular disease was estimated to have a median value greater than six, indicating that the hospitalization risk of COVID-19 patients with cardiovascular disease was six times greater than that of COVID-19 patients without any comorbidities. The hospitalization risk for COVID-19 patients also increased with chronic respiratory disease (asthma and COPD), hypertension, and obesity. These comorbidities were selected for this study since they were documented in both the COVID-NET and the New York datasets. Our preliminary exploration with the COVID-NET dataset also indicated elevated risks for people with autoimmune diseases, immune suppression conditions, and renal diseases (data not shown). We encourage other researchers to apply our Bayesian model to their own investigations.

One limitation with our current analysis is that our estimated hospitalization risk for patients with a comorbidity of interest (e.g., diabetes) may be confounded with other comorbidities in the same patient (e.g., hypertension). This limitation is attributed to our lack of access to the necessary data rather than our Bayesian approach *per se*. In this study, we relied upon two published summary statistics (i.e., COVID-NET and New York), which did not include the details of joint comorbidities in their publications^3,5^. For researchers who can access the complete medical records (instead of just summary statistics), they would be able to obtain the frequency of joint comorbidities (e.g., number of COVID-19 hospitalized patients with both diabetes and hypertension). Then, our Bayesian approach could be applied directly to estimate the hospitalization risk for such joint comorbidities. Specifically, instead of using the frequency of a particular comorbidity (e.g., diabetes), our model would use the frequency of the joint comorbidities (e.g., diabetes and hypertension). In addition, the joint prevalence of comorbidities (e.g., diabetes and hypertension) would be used as informative priors.

The only biological assumption in our Bayesian model is that people in the general population, regardless of the status of their comorbidities, could be equally infected by the SARS-CoV-2 virus (no assumptions on the severity of the symptoms after infection were made). The assumption of equal chance of infection by the virus is based on the rationale that SARS-CoV-2 is a newly emerged virus to the human population; thus, nobody is particularly immune to the virus. For example, it has been reported that viral loads were similar in asymptomatic and symptomatic patients^6^, and young children and adults were both similarly infected by the virus^7^. If future research shows that people with specified comorbidities do have a different chance of being infected, our Bayesian approach can be modified to accommodate that difference (e.g., using a scaling factor in our model to reflect the differential degree of infection probability).

## Materials and Methods

### Bayesian modeling

The overview of our Bayesian model is depicted in the figure 2. We classify people in the general population into three categories: (1) *h* - the people without any known comorbidities, (2) *c* - the people with a particular comorbidity of interest (e.g., diabetes), who may or may not carry other types of comorbidities, and (3) *o* - the people with some other types of comorbidities excluding *c* (e.g., any other types of comorbidities excluding hypertension). Let *N*_*h*_, *N*_*c*_, and *N*_*o*_ denote the number of hospitalized COVID-19 patients for these three categories, respectively. Let *N* denote the total number of COVID-19 patients in the hospital, i.e., *N* = *N*_*h*_ + *N*_*c*_ + *N*_*o*_. Then the vector (*N*_*h*_, *N*_*c*_, *N*_*o*_) is a multinomial random variable^8^:

**Figure 2.**
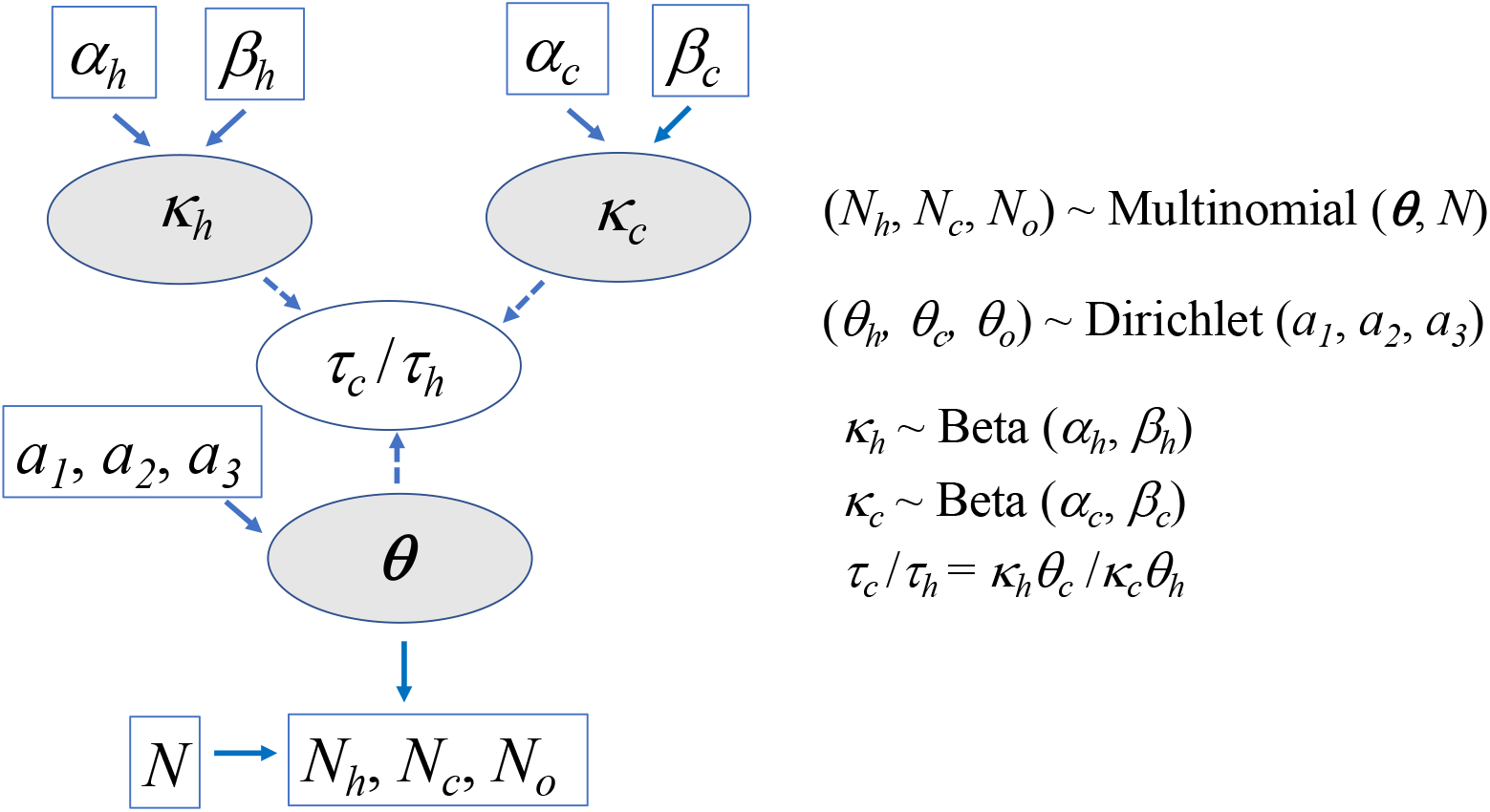
Graphic representation of the Bayesian model. On the left, the boxes represent constants for either fixed parameter values (*α*_*h*_, *β*_*h*_, *α*_*c*_, *β*_*c*_, *a*_*1*_, *a*_*2*_, *a*_*3*_) for priors or observed data (*N, N*_*h*_, *N*_*c*_, *N*_*o*_). The grey ovals (*κ*_*h*_, *κ*_*c*_, *θ*_*h*_) represent stochastic nodes. The white oval (*τ*_*c*_ /*τ*_*h*_) represents a deterministic node. The solid arrows represent stochastic dependence. The dashed arrows represent logical dependence. The corresponding stochastic and deterministic expressions are depicted on the right.

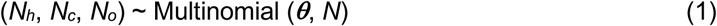

where ***θ*** denotes a vector of unobserved multinomial probabilities (*θ*_*h*_, *θ*_*c*_, *θ*_*o*_) corresponding to *N*_*h*_, *N*_*c*_, and *N*_*o*_, respectively.

Let *τ*_*h*_, *τ*_*c*_, and *τ*_*o*_ denote the unknown probabilities of hospitalization for people infected by SARS-CoV-2 virus in the category of *h, c*, and *o*, respectively, in the general population. For example, if 50,000 out of a total of 200,000 infected people with a particular comorbidity of interest (i.e., in the category of *c*) were eventually hospitalized, *τ*_*c*_ would be equal to 0.25 (i.e., 50,000/200,000). We define *τ*_*c*_/*τ*_*h*_ to be the risk ratio of the probability of hospitalization for the infected people with a particular comorbidity of interest *c* (e.g., diabetes) versus the probability of hospitalization for the infected people without any comorbidities. It’s important to note that *τ*_*h*_, *τ*_*c*_, and *τ*_*o*_ are different from *θ*_*h*_, *θ*_*c*_, and *θ*_*o*_. However, we derived an algebraic relationship between *τ*_*c*_/*τ*_*h*_ and *θ*_*c*_/*θ*_*h*_, as shown below.

Let *κ*_*h*_ and *κ*_*c*_ denote the proportion of people without any medical conditions and the people with a comorbidity of interest *c* (e.g., diabetes), respectively, in the general population of size *N*_*pop*_. Let *ρ*_*h*_, *ρ*_*c*_, and *ρ*_*o*_ denote the probabilities of being infected by SARS-CoV-2 virus for people in the categories of *h, c*, and *o*, respectively. Then, ***θ*** can be expressed as follows:

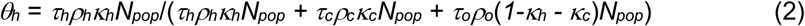

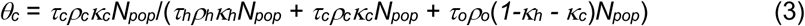

In Eq. (2), the numerator (*τ*_*h*_*ρ*_*h*_*κ*_*h*_*N*_*pop*_) corresponds to the expected number of hospitalized COVID-19 patients without any comorbidities. Specifically, out of the general population of size *N*_*pop*_, *κ*_*h*_*N*_*pop*_ is the expected number of people without any comorbidities given the definition of *κ*_*h*_; *ρ*_*h*_*κ*_*h*_*N*_*pop*_ is the expected number of people without any comorbidities infected by the virus given the definition of *ρ*_*h*_; *τ*_*h*_*ρ*_*h*_*κ*_*c*_*N*_*pop*_ is the expected number of infected people without any comorbidities who were hospitalized given the definition of *τ*_*h*_. Similarly, in Eq. (3), the numerator (*τ*_*c*_*ρ*_*c*_*κ*_*c*_*N*_*pop*_) corresponds to the expected number of hospitalized COVID-19 patients with the comorbidity of interest *c*. In addition, the term *τ*_*o*_*ρ*_*o*_(*1-κ*_*h*_ *-κ*_*c*_)*N*_*pop*_ in the denominator of both Eq. (2) and (3) corresponds to the expected number of hospitalized COVID-19 patients with some other types of comorbidities excluding *c*. Therefore, the denominator in both Eq. (2) and (3) corresponds to the expected total number of hospitalized COVID-19 patients, regardless of the status of their comorbidities.

If we assume that people in the general population, regardless of the status of their comorbidities, have an equal chance of being infected by SARS-CoV-2 virus (see Discussion), then *ρ*_*h*_ = *ρ*_*c*_ = *ρ*_*o*_ and the Eq. (2) and (3) can be simplified by canceling out *ρ*_*h*_, *ρ*_*c*_, *ρ*_*o*_, and *N*_*pop*_ from both the numerator and denominator. The simplified Eq. (2) and (3) are shown as follows:

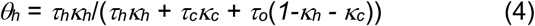

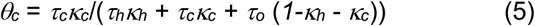

Dividing Eq. (5) by Eq. (4), we obtain the expression of *τ*_*c*_/*τ*_*h*_ as follows:

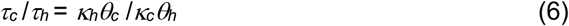

To estimate the posterior probability of *τ* /*τ*_*h*,_ we need to sample from the following posterior distribution:

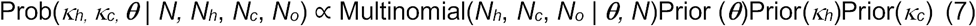

To specify the prior distribution for ***θ***, we chose a Dirichlet distribution^8^ as it is commonly used as the conjugate prior of the multinomial likelihood described in Eq. (1).

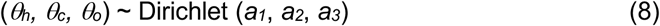

where *a*_*1*_, *a*_*2*_, *a*_*3*_ correspond to the shape parameters of Dirichlet distribution. For this study, we set *a*_*1*_ = *a*_*2*_ = *a*_*3*_ = 1 to set a uniform prior, although those values can be adjusted to more accurately reflect the probabilities of hospitalization for each category of patients if more data becomes available in the future.

To specify the prior distributions for *κ*_*h*_ and *κ*_*c*_, we chose beta distributions as they are commonly used to model proportions^8^.

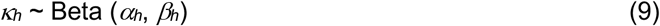

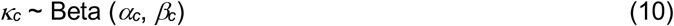

where *α*_*h*_, *β*_*h*_, *α*_*c*_, and *β*_*c*_ denote shape parameters of the corresponding beta distributions. Using the method of moments^8^, these parameters can be expressed as follows:

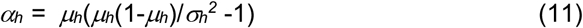

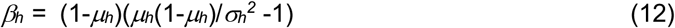

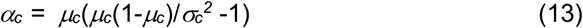

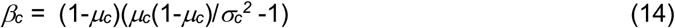

where *μ_h_* and *σ_h_^2^*, and *μ_c_* and *σ_c_^2^* represent the mean and variance of the proportions of the healthy people and people with the comorbidity of interest *c*, respectively, in the general population.

Then, by plugging in the priors in Eq. (7), the posterior distribution becomes the following:

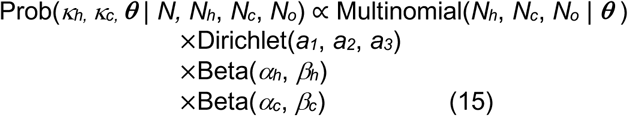

In summary, the foundation of our approach is based on our derived algebraic relationship (Eq. 6) between the quantity of *τ*_*c*_/*τ*_*h*_ (the risk ratio) and the quantities of *κ*_*h*,_ *κ*_*c*,_ and ***θ***. Using a uniform Dirichlet distribution, ***θ*** is modeled by a noninformative prior; ***θ*** is related to the observed data (*N, N*_*h*_, *N*_*c*_, and *N*_*o*_) in hospitalized COVID-19 patients through the multinomial likelihood as described in Eq. (1). *κ*_*h*_ and *κ*_*c*_ are modeled by informative priors using beta distributions whose shape parameters were expressed using the published prevalence rates for comorbidities in the general population. Through sampling from the posterior distribution of *κ*_*h*,_ *κ*_*c*,_ and ***θ***, we were able to estimate the posterior distribution of *τ*_*c*_/*τ*_*h*_ as a derived quantity of *κ*_*h*_*θ*_*c*_ /*κ*_*c*_*θ*_*h*_.

We used WinBUGS^9^ (version 1.4.3) to implement the above models. The posterior distributions of risk ratios for different comorbidities were estimated with the Markov Chain Monte Carlo (MCMC) sampling strategy implemented in WinBUGS^10^ using the following parameters: the number of chains of four, the number of total iterations of 100,000, burn-in of 10,000, and thinning of 4. Convergence and autocorrelations were evaluated with trace/history and autocorrelation plots. Multiple initial values were applied for MCMC sampling.

### Comorbidity data for hospitalized COVID-19 patients

For the above Bayesian approach, the following two types of data are required: (1) the frequency of the comorbidity of interest (e.g., diabetes) in COVID-19 patients in hospitals, and (2) the prevalence of the comorbidity in the general population. For the comorbidity frequency of hospitalized COVID-19 patients, we used a large-scale dataset, available at COVID-NET^5^, collected from 154 acute care hospitals in 74 counties in 13 states in U.S. from March 1 to May 2, 2020. Among a total of 2491 hospitalized adult patients with laboratory-confirmed COVID-19 in this COVID-NET dataset, 314 had asthma, 266 had COPD, 859 had cardiovascular diseases, 819 had diabetes, 1154 were obese, 1428 had hypertension, and 336 had no known medical conditions. Besides the COVID-NET dataset, we also used a published dataset from the state of New York^3^ collected from 12 hospitals in New York City, Long Island, and Westchester County from March 1 to April 4, 2020. Among a total of 5700 hospitalized COVID-19 patents in this New York dataset, 479 had asthma, 287 had COPD, 966 had cardiovascular diseases, 1808 had diabetes, 1737 were obese, 3026 had hypertension, and 350 had no known medical conditions. In both the COVID-NET and New York datasets, cardiovascular disease referred to coronary artery disease and congestive heart failure.

For the prevalence of comorbidities in general U.S. adult population, the following estimates (mean ± standard error) by the U.S. public health government agencies were used: asthma (7.7%±0.22%)^11^, cardiovascular disease (5.6%±0.14%)^12^, COPD (5.9%±0.051%)^13^, diabetes (13%±5.6%)^14^, obesity (42.4%±1.8%)^15^, and hypertension (49.1%±1.5%)^16^. The proportion of healthy adults in the U.S. who have no medical conditions was estimated to be 12.2% (95% CI: 10.9–13.6)^17^.

## Data Availability

All the relevant data are described in the manuscript and the supplementary material

## Acknowledgments

We thank Drs. Lindsay Kim and Gayle E. Langley at the U.S. Centers for Disease Control and Prevention for providing the COVID-NET dataset for our analysis.

